# Host behaviour driven by awareness of infection risk amplifies the chance of superspreading events

**DOI:** 10.1101/2023.08.25.23294423

**Authors:** Kris V Parag, Robin N Thompson

## Abstract

We demonstrate that heterogeneity in the perceived risks associated with infection within host populations amplifies the chances of superspreading during the crucial early stages of an epidemic. Under this behavioural model, individuals less concerned about the dangers from infection are more likely to be infected and attend larger-sized (riskier) events. For directly transmitted diseases such as COVID-19, this leads to infections being introduced at rates above the population prevalence to the events most conducive to superspreading. We develop an interpretable computational framework for evaluating within-event risks and derive a small-scale reproduction number measuring how the infections generated at an event depend on transmission heterogeneities and the number of introductions. This quantifies how event-scale patterns relate to population-level characteristics and generalises previous frameworks. As event duration and size grow, our reproduction number converges to the basic reproduction number. We illustrate that even moderate levels of heterogeneity in the perceived risks from infection substantially increase the likelihood of disproportionately large clusters of infections occurring at larger events, despite fixed overall disease prevalence. We show why collecting data linking host behaviour and event attendance is essential for accurately assessing the risk posed by an invading pathogen in the emerging stages of an outbreak.

## Introduction

The prediction and prevention of superspreading events, which are characterised by primary infected individuals generating disproportionately large numbers of secondary infections [1], is a central challenge in infectious disease epidemiology. For acute, directly communicable diseases such as COVID-19, SARS and Ebola virus disease, superspreading is a major driver of transmission that leads to less frequent but more explosive outbreaks than we might expect under more classical models that neglect the substantial variability in secondary infections generated by infected hosts [2]. During early or emergent stages of a potential epidemic, when there are limited immunity levels in the host population and transmission dynamics are inherently stochastic, superspreading events have been found responsible for spurring both the initial growth and eventual persistence of epidemics and for limiting the effectiveness of non-pharmaceutical interventions [1,3–5].

Consequently, identifying the main factors that underly the risk of superspreading is crucial for effective disease management [4]. Many of these factors are known, with heterogeneities in (i) host characteristics (e.g., susceptibility, infectiousness and contact patterns), (ii) pathogen biology (e.g., transmission routes and viral loads), (iii) environmental effects (e.g., ventilation and gathering size) and (iv) host behaviours (e.g., social customs and intervention adherence) all contributing to the risk of superspreading [3,4,6–8]. However, incorporating these factors in parsimonious modelling frameworks can be difficult because the mechanisms linking them to superspreading are still not fully understood. This is particularly the case for factors (iii) and (iv), with recurrent calls for more comprehensive data collection to help study the relationships among behavioural, environmental and epidemiological trends [9–11]. Here we explore how a key feature of host behaviour can shape the likelihood of superspreading and provide a mathematical demonstration of the benefits of collecting and analysing more data to elucidate the links between human behaviour and infectious disease epidemics.

We consider how heterogeneity in perceptions of the risk associated with infection throughout a host population impact heterogeneity in the transmission of new infections in the early stages of an epidemic. Risk awareness is a documented phenomenon in which individuals engage in self-protective behaviours in response to the perceived health, economic and other dangers of acquiring infection. The exact relationship between risk-perception and self-protection in a population exists on a spectrum spanning more risk-averse individuals to those with larger risk appetites (e.g., risk deniers) [12–14]. During an epidemic of a directly transmitted pathogen, risk-averse individuals may reduce their number of contacts by limiting their socialising and mobility, while those with larger risk appetites may increase their relative infection exposure (e.g., by hosting unregulated gatherings when infections are rising) [15]. Risk awareness can improve intervention efficacy (e.g., reducing mobility) or negate it (e.g., via deliberate non-compliance) and substantially change outbreak amplitudes and durations [11,16–19].

Despite its importance, the interplay between risk awareness and superspreading risk has not been studied in detail, with most research focussing on pathogen and host characteristics instead of exploring behavioural patterns. We study this interplay under a simple but plausible hypothesis – that more risk-averse hosts are more likely to avoid events of larger size, due to the perception of heightened infection risk at those events [14,19]. Here, events are short-term gatherings (e.g., parties) so that only one generation of infection is possible. While data directly linking risk perception to event attendance are unavailable, it is known that individuals modify their behaviour in response to population-level prevalence and that the variability in individuals’ level of acceptable risk relative to this prevalence baseline correlates well with the extent to which contacts are reduced during epidemics [12–14,20,21]. A likely pathway for reducing exposure to invading pathogens is by limiting attendance at riskier (voluntary) events. This logic underlies our hypothesis and subsequent analysis.

This awareness mechanism implies, for a fixed population prevalence, that larger events (e.g., concerts, sports matches) are more likely to be attended by individuals who are less risk averse. Since there is limited infection-induced immunity in the host population during early epidemic stages, these individuals are also more likely to be infected. This stems from observations that those with larger numbers of contacts have elevated chances of acquiring infection, and these individuals are also likely to have larger risk appetites (more risk-averse individuals tend to reduce contacts) [22]. We posit that this coupling between behaviour and environment (i.e., modification of event attendance due to risk perception and event size) may amplify the chances of superspreading occurring at larger events, which have the capacity to support excessive numbers of infections. To test this hypothesis, we develop a framework to model the number of infections *y* generated at an event of size *n*, given that *x* initially infected individuals attend that event. This yields a small-scale reproduction number that extends recent approaches [23–25] to understanding within-event transmission in three directions.

First, we explicitly model the transmission-reducing effects of finite numbers of susceptible individuals (*n* − *x*) and imported infections (*x*) and at the event. As event size and duration grow, these finite size effects become less important and our small-scale reproduction number converges to *R*_0_, the popular basic reproduction number. Second, we embed heterogeneity in transmission at the event within our small-scale reproduction number by allowing variations in secondary infections that are controlled by a dispersion parameter, *k*. This is a within-event version of the seminal model of superspreading [1,5,26] and includes the broad influence of factors (i)-(ii) described above. Third, we account for how *x* changes (stochastically) with *n*. This considers factors (iii)-(iv) and depends on the prevalence of infection in the population as well as the size-biased importation rate of infections into the event, *ϵ*(*n*), which is influenced by the spectrum of risk appetites about that prevalence.

The functional dependence of *ϵ* on *n* serves as a parsimonious model of risk awareness and allows us to assess how host behaviour shapes the risk of superspreading (e.g., if *ϵ*(*n*) is an increasing function of *n*, then this implies a higher infection import rate into larger events). We explore our central hypothesis by comparing the relative and combined impact of *ϵ*(*n*) and *k* on the tail probability of observing a disproportionately large number of secondary infections *y* at an event. We demonstrate, for a fixed overall import rate (equalling the wider population prevalence), that risk awareness can substantially amplify the chances of superspreading at a large event, compared to the scenario in which all individuals attending the large event are assumed to have similar perceptions of infection risk. This pattern holds regardless of *k* and, in some instances, we find that the increase in superspreading risk from risk-aware behaviour outweighs that from inherent transmission heterogeneity.

## Methods

### Event reproduction numbers including import risk and transmission heterogeneity

We develop a framework for quantifying the risk of acquiring infection at an event (e.g., a party, concert, sports match), based on a small-scale (within-event) reproduction number. We detail this below but also sketch the main steps of our methodology and list key notation in **Fig *1***. An event is defined as a short-term grouping of *n* people and we allow 0 ≤ *x* ≤ *n* of the individuals attending the event to be infectious. Initially, there are *x* introductions (i.e., imported infections) at this event and *n* − *x* susceptible hosts. We assume no prior immunity in the population and let **P**(*y*|*n*) be the probability of 0 ≤ *y* ≤ *n* − *x* new infections being generated at that event as we describe in **Eq. (1)**.

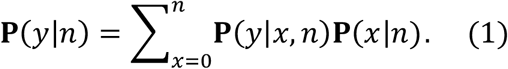

This depends on **P**(*y*|*x, n*), the probability of *y* new infections occurring given the *x* infectious individuals initially (for events of size *n*) and the prior probability of those *x* imports, **P**(*x*|*n*). We define the small-scale reproduction number for this event as 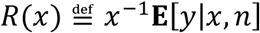, with the expected number of infections generated by *x* imports denoted by **E**[*y*|*x, n*]. We expand this to obtain **Eq. (2)** below.

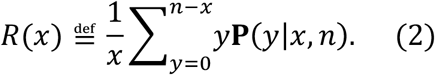

Here *R*(*x*) measures the expected number of new infections generated by each import when there are *x* imports in total. Although intuitive, this reproduction number formulation is novel. A central idea of this study is the importance of **P**(*x*|*n*) and its dependence on event size *n*. Earlier work assumed that **P**(*x*|*n*) depends solely on the prevalence of the infection in the population [25], neglecting how heterogeneities in human behaviour may affect the number of imported cases at a given event of size *n*. To our knowledge, alternative models for **P**(*x*|*n*) informed by human behaviour and the influence of this behaviour on the number of infections generated at the event have not been explored. The heterogeneity in host behaviour that we consider relates to the spectrum of risk appetites i.e., the fact that different individuals perceive different infection risks associated with attending an event of size *n*. This spectrum alters the rate of importing infections into events, relative to the prevalence, and so modulates **P**(*x*|*n*).

**Fig 1:**
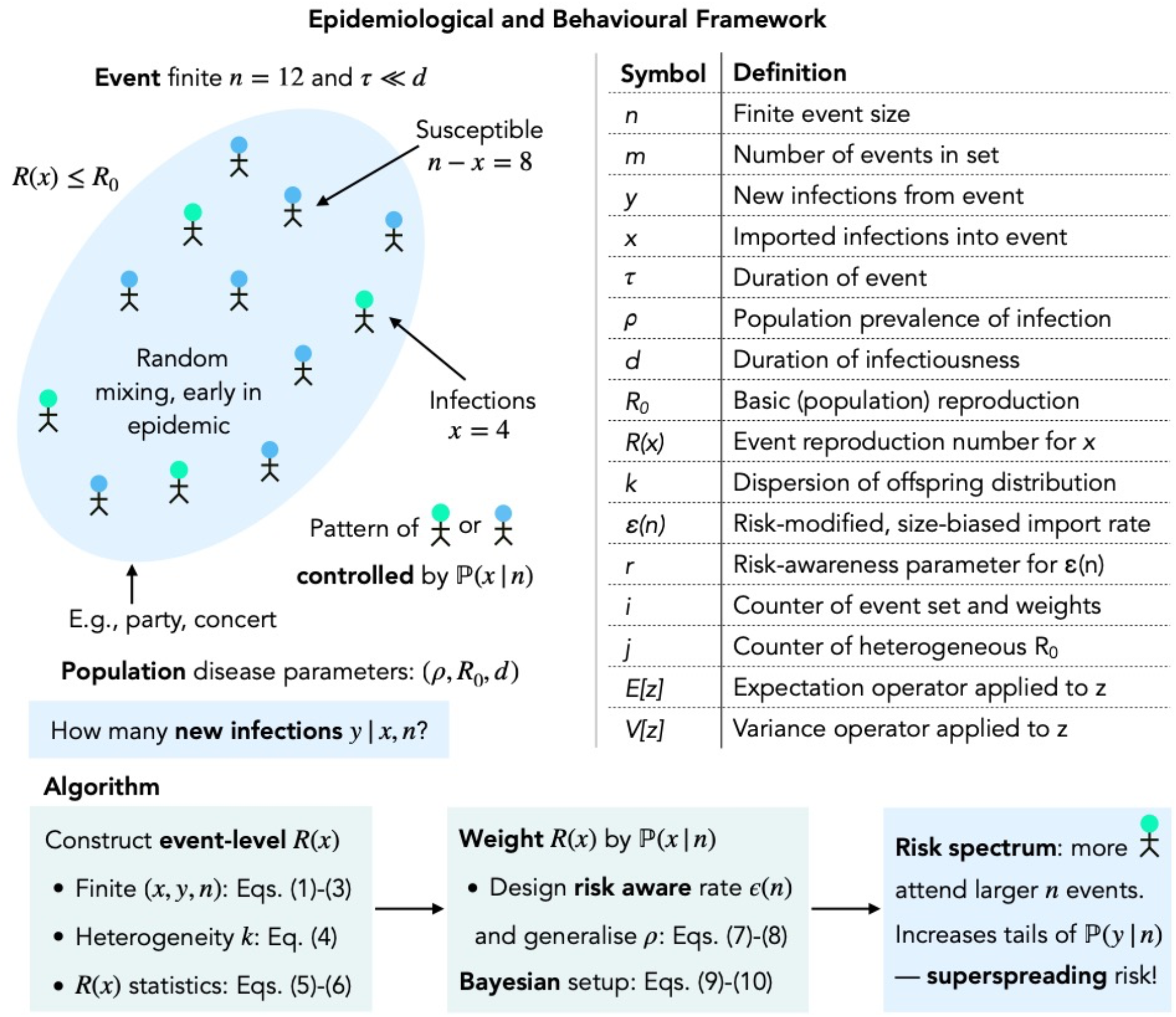
Modelling framework for event-level transmission subject to risk awareness. We outline the central steps and define the main notation underlying our proposed framework for modelling transmission patterns at small events. We refer to equations defined in the Methods. This framework accounts for how heterogeneity in infection risk perception among individuals modulates the number of imported cases *x* at an event of size *n* and hence contributes to the secondary infections generated at that event *y*.

Our event or small-scale reproduction number also generalises prior research by including the effects of finite *x* and *n*. Since only one generation of infection can occur at an event, this finite initial condition can strongly shape clustering patterns, underscoring the value of modelling **P**(*x*|*n*). The original event reproduction number [24] considers a single imported infection and relates to (but is not the same as) our *R*(1), which we later show is always an upper bound for *R*(*x*). By extending the event reproduction number definition, we model the influence of **P**(*x*|*n*) on the risk of acquiring infection at any event directly. As we explain below, *R*(*x*) also embeds heterogeneity in transmission from both host characteristics as well as pathogen biology [1] and is explicitly related to the population-level basic reproduction number, *R*_0_ [27].

To convert **Eqs. (1)-(2)** into a computable form, we draw on characteristics of both the event and disease. We denote the (frequency-dependent) transmission rate as *β* and the expected duration of an individual infection as *d*, so that *R*_0_ = *βd*. We then consider an event that lasts for time τ, which is assumed to be substantially shorter than *d*, so that infectiousness outlasts the event and at most one generation of infection is possible at the event. We also assume that the event is closed i.e., for any specific event, *n* takes a constant value. The split of *n* into *x* and *n* − *x* completely defines the epidemiologically important states for the event

If there is only one infected individual at the start of the event, then the probability that any susceptible host gets infected is the secondary attack rate (SAR), 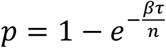, making the standard assumption that the times to infection are exponentially distributed. When there are *s* susceptible individuals, then **E**[*y*|1, *n*] = *sp*. While this assumes that all the susceptible individuals are exposed to all infectious ones at the event, we can model more realistic contact networks as in [27] by modifying *s* to be the subset of susceptible hosts likely to be exposed to each infection (this connects network and random mixing models).

We generalise this approach in three main directions. First, we model the effect of variability in the number of imported infections. If there are *x* imports to the event, then the SAR becomes 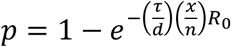 with 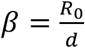. Since there are initially *s* = *n* − *x* susceptible individuals, the expected number of infections generated at the event is **E**[*y*|*x, n*] = (*n* − *x*)*p*. The leads to the event reproduction number *R*(*x*) in **Eq. (3)** below. Note that *R*(0) = *R*(*n*) = 0.

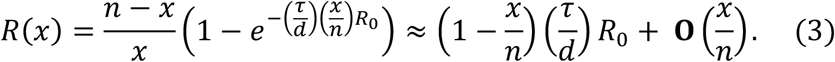

This formulation has interesting limiting behaviour at various *x*. As the number of susceptibles grows in excess of imports i.e.,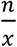 increases, the 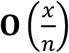 terms in the Taylor series approximation of *R*(*x*) in **Eq. (3)** become negligible. As *n* becomes large, we find 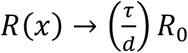. If the event lasts for the duration of infectiousness (τ = *d*), then *R*(*x*) → *R*_0_. This convergence makes sense since our formulation is equivalent to a finite or small-scale version of random mixing.

Second, we expand this model to include realistic heterogeneity from host characteristics and pathogen biology. It is unlikely that every infectious individual has the same transmissibility and we expect substantial variations in the numbers of infections generated by each infected individual [1,28]. We therefore allow *R*_0_ to have some distribution from which every import is randomly sampled and let 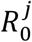 indicate the sample for the *j*^th^ of the *x* imports at the event. This heterogeneous version of *R*(*x*) is in **Eq. (4)**, with expected number of infections **E**[*y*|*x, n*] = *xR*(*x*). Note that **Eq. (4)** is of the form 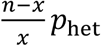, with *p*_het._ as a heterogeneous SAR.

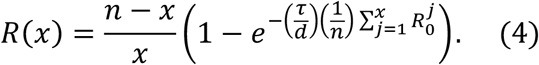

We compute the mean of *R*(*x*) across the transmission heterogeneity for *x* infectious imports in **Eq. (5)**, with **E**_het._ indicating expectation about the distributions of the 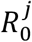 and **M**_b_(*a*) as the moment generating function about *b* evaluated at *a*. As the transmissibility of the *x* imported infections are independently sampled, 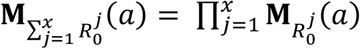. This reduces to 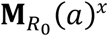 if samples are identically distributed. The expected number of infections under this model as a function of *x* is **E**_het._[**E**[*y*|*x, n*]] = *x***E**_het_[*R*(*x*)] with **E**_het_[*R*(*x*)] from **Eq. (5)**.

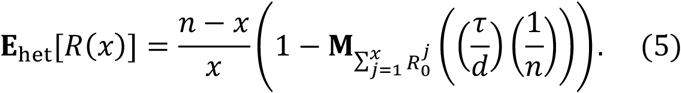

Following [28], we evaluate the variance around *R*(*x*) as **V**_het_[*R*(*x*)] with 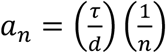 in **Eq.(6)**. This involves expanding **E**_het_[*R*(*x*)^2^] – **E**_het_[*R*(*x*)]^2^ and applying properties of **M**_b_(*a*). The variance on the expected number of infections is **V**_het_[**E**[*y*|*x, n*]] = *x*^2^**V**_het_[*R*(*x*)]. All of these statistics remain valid for any model of transmission heterogeneity but we derive analytic relations under the most widely used model of [1] in the subsequent section.

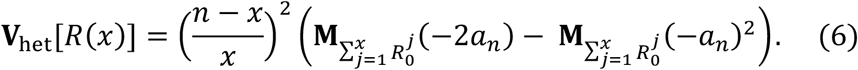

Third, we examine how the likelihood of finding that *x* infectious individuals have attended the event impacts the above quantities. This involves evaluating how **P**(*x*|*n*) weights the formulae in **Eqs. (4)-(6)**. This weighting may be random, depend on behavioural preferences as we posit in the next section (i.e., risk awareness) or be assigned using other rules. We propose that a more informative measure of the risk of acquiring infection from an event of size *n* and duration τ is the import-weighted event reproduction number *R*_imp_ as in **Eq. (7)** below.

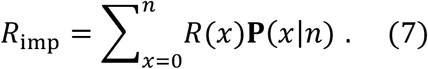

While *R*_imp_ averages over the possible numbers of imports, it is still a random variable with samples taken from the distribution controlling transmission heterogeneity. Accordingly, it has statistics **E**_het_[*R*_imp_] and **V**_het_[*R*_imp_] that we compute by summing and weighting **E**_het_[*R*(*x*)] and **V**_het_[*R*(*x*)] by **P**(*x*|*n*) and **P**(*x*|*n*)^2^ respectively. The expected number of new infections **E**_imp_[*y*|*n*] that is associated with *R*_imp_ follows as in **Eq. (8)**.

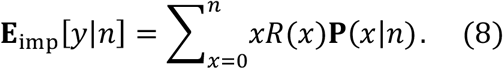

Similarly, we obtain the heterogeneous statistics **E**_het_ [**E**_imp_[*y*|*n*]] and **V**_het_ [**E**_imp_[*y*|*n*]] but the quantities being weighted by **P**(*x*|*n*) and **P**(*x*|*n*)^2^ are now, respectively, *x***E**_het_[*R*(*x*)] and *x*^2^**V**_het_[*R*(*x*)]. These all proceed from the properties of expectations and variances applied to a linear weighted sum with independent terms.

### Statistical models for event reproduction numbers and importation patterns

Having outlined measures of infection risk in **Eqs. (7)-(8)**, we build into our framework some likely approaches for integrating transmission heterogeneities and import patterns (including when those imported infections are risk-sensitive). This allows us to parsimoniously model traditional and behavioural drivers of superspreading. Additionally, we incorporate process stochasticity and provide a full Bayesian formulation for our framework. We start by including the seminal heterogeneity model of [1], which describes individual variations in transmissibility via a gamma distribution with dispersion *k* and mean *R*_0_. We write this as 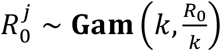 with **Gam** as a shape-scale parameterised gamma distribution. Using scaling and summing properties of these gamma variables, we hence obtain 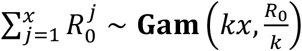.

This assumes that samples of the basic reproduction number of individuals are independent and identically distributed and lets us analytically evaluate the moment generating function as 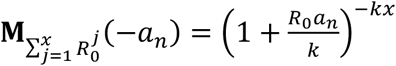. We substitute this into **Eqs. (4)-(6)** to precisely compute the mean and variance of the infections and event reproduction number conditional on a total of *x* introductions as detailed above. We can relax the assumption that the 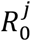 are independent and identically distributed by instead sampling them from different distributions or by applying alternative dispersion models [28]. The heterogeneous 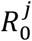 constitute a major and traditionally modelled source of stochasticity underpinning the risk metrics we propose in **Eqs. (7)-(8)**.

A less studied source of stochasticity is variability in the probability that infectious individuals attend the event. Previous work [25] has treated this deterministically, setting the probability or rate that an attending individual is infected as equal to the population prevalence *ρ* (or *ρ* adjusted by an exposure factor when it is known that the event draws individuals who are less or more likely to be infected). This is modelled as *x ∼* **Bin**(*n, ρ*), with **Bin** indicating a binomial distribution. We generalise this under our behavioural hypothesis. We posit, for a fixed overall importation level, that this import probability increases with *n*. This models risk awareness, in which risk-averse individuals who are less likely to be infected avoid larger events, or equally the individuals attending larger sized events are less risk-averse and more likely to be infected. Risk appetites may also depend on event duration τ, but we do not explore this here.

We model event size bias using sorted Dirichlet weightings. We consider *m* events, the *i*^th^ of which has size *n*_*i*_ and import rate *ϵ*(*n*_*i*_). Sizes sequentially span all integers from *n*_*min*_ to *n*_*max*_ uniquely (i.e., *n*_*max*_ = *n*_*min*_ + *m* − 1) but we can relax this to include any distribution across event sizes of interest. We fix the total importation rate across all *m* events. This constrain 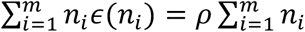, conserving the total number of infections introduced across events so that the mean importation rate equals *ρ*. We enforce this constraint to allow fair comparison between the conventional model, in which all imports occur with rate *ρ*, and our size-biased variations, which describe how variability in perceived risk by hosts affect their attendance at events. This constraint causes some event sizes to have importation rates above and others below *ρ* and allows us to model a spectrum of risk appetites about the baseline prevalence. This variability in risk perception aligns well with trends found in behavioural surveys [12–14].

The *ϵ*(*n*_*i*_) values encode our event size biases. We construct them by first sampling a set of random weights {*w*_*i*_} from a symmetrical Dirichlet distribution i.e., {*w*_*i*_} *∼* **Dir**(*r*) with *r* as a shape parameter applied to every *w*_*i*_ and 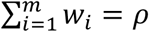. The {*w*_*i*_} set spans all *m* weights with smaller values of *r* leading to more skewed weightings. At very large *r*, 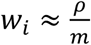 for all *i*. To model risk-awareness, where we expect that less risk-averse individuals are more likely to attend larger *n*_*i*_ events relative to more risk-averse individuals, we sort the {*w*_*i*_} in ascending order so that *w*_*i*_ increases with *n*_*i*_. We replicate this procedure across many runs to include variability from the Dirichlet distribution. For every sampled, sorted {*w*_*i*_}, we define 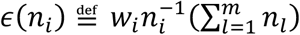. This satisfies our benchmarking constraint and parsimoniously models the spectrum of risk appetites across the host population.

We conceptualise this constraint by observing that in the conventional model **E**[*x*_*i*_] = *n*_*i*_*ρ* for the *i*^th^ event so that 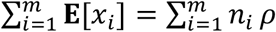 imports occur into all *m* events on average. In our risk-aware model **E**[*x*_*i*_] = *n*_*i*_*ϵ*(*n*_*i*_) and so we chose the *ϵ*(*n*_*i*_) to ensure 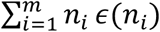 equals the 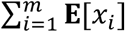 from the conventional model. However, we can relax this constraint to describe when risk awareness itself changes the prevalence (provided we use updated values at time snapshots) and we can generalise the model to allow *r* to also be size dependent (i.e., *r*(*n*_*i*_)). In summary, we generate import rates that are size biased (with this bias accounting for risk awareness) and use a single parameter *r* to set the strength of this behavioural effect.

Integrating the above models for heterogeneity and importation, we complete our algorithm (see **Fig *1***) for sampling import weighted distributions of event size risk using **Eqs. (1)-(2)**. We formulate this in **Eqs. (9)-(10)** with semi-colons discriminating between probabilities that we evaluate from a distribution and parameters specifying that distribution. For convenience, we use 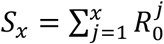 for denoting heterogeneous samples and *ϵ*(*n*) for general size bias.

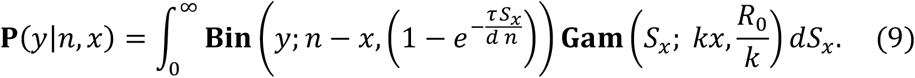

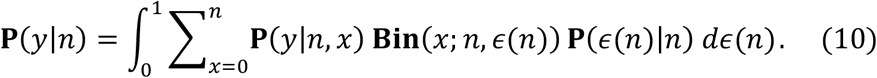

We use the probability distributions in **Eqs. (9)-(10)** together with the definitions of **Eqs. (1)-(2)** to compute the measures of event risk that we propose in **Eqs. (7)-(8)**. These marginalise over the distributions of import rate and transmission heterogeneity, which are degenerate when *ϵ*(*n*) is constant for all *n* or all 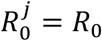, respectively. In the Results, we examine the properties of our computational framework and apply it to explore how behaviour affects superspreading. Our framework is freely available at: https://github.com/kpzoo/smallscaleR.

## Results

In the Methods, we developed a framework to assess the risk of acquiring infection at an event by deriving a small-scale reproduction number and the expected number of infections that will occur at the event. Both measures depend on the levels of heterogeneity in transmission and variability in the rate at which infectious individuals are likely to attend the event (i.e., imports). Here we examine the influence of these two key factors in determining outbreak patterns.

### Superspreading risk depends on importations and dispersion

Much research has investigated how heterogeneity in transmission can cause superspreading and hence increase the number of infections likely to result from a gathering or event [1,23]. Specifically, there has been study of how the dispersion parameter *k* modulates the risk of superspreading events [26,28,29]. Generally, smaller values of *k* < 1 are predictive of larger transmission heterogeneity and superspreading risk. However, the influence of the number of importations *x* at an event of size *n* has received relatively little attention. We examine this by computing the statistics derived in **Eqs. (4)-(6)**, in which we defined the reproduction number *R*(*x*) as a function of the imports and the resulting number of expected infections **E**[*y*|*x, n*].

We consider an event of size *n* = 30 over a range of dispersions 0.1 ≤ *k* ≤ 10 with a large-scale limit (see **Eq. (3)**) of 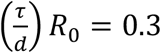. We sample *R*(*x*) and **E**[*y*|*x, n*] from heterogeneous gamma distributions describing the transmissibility of the sum of all imported infections (see Methods) and compute statistics from these samples using **Eqs. (5)-(6)**. We plot these results in **Fig *2*** to explore the properties of these statistics. Interestingly, we find *R*(*x*) is a decreasing function of *x*, even though every *R*(*x*) has the same limit of 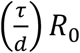. The single import scenario of *R*(1) relates to the event reproduction number proposed in [24]. If we assume, as in several branching process models, that all imports have reproduction number *R*(1) instead of *R*(*x*), then **E**[*y*|*x, n*] and the risk of acquiring infection at the event may be notably overestimated.

**Fig 2:**
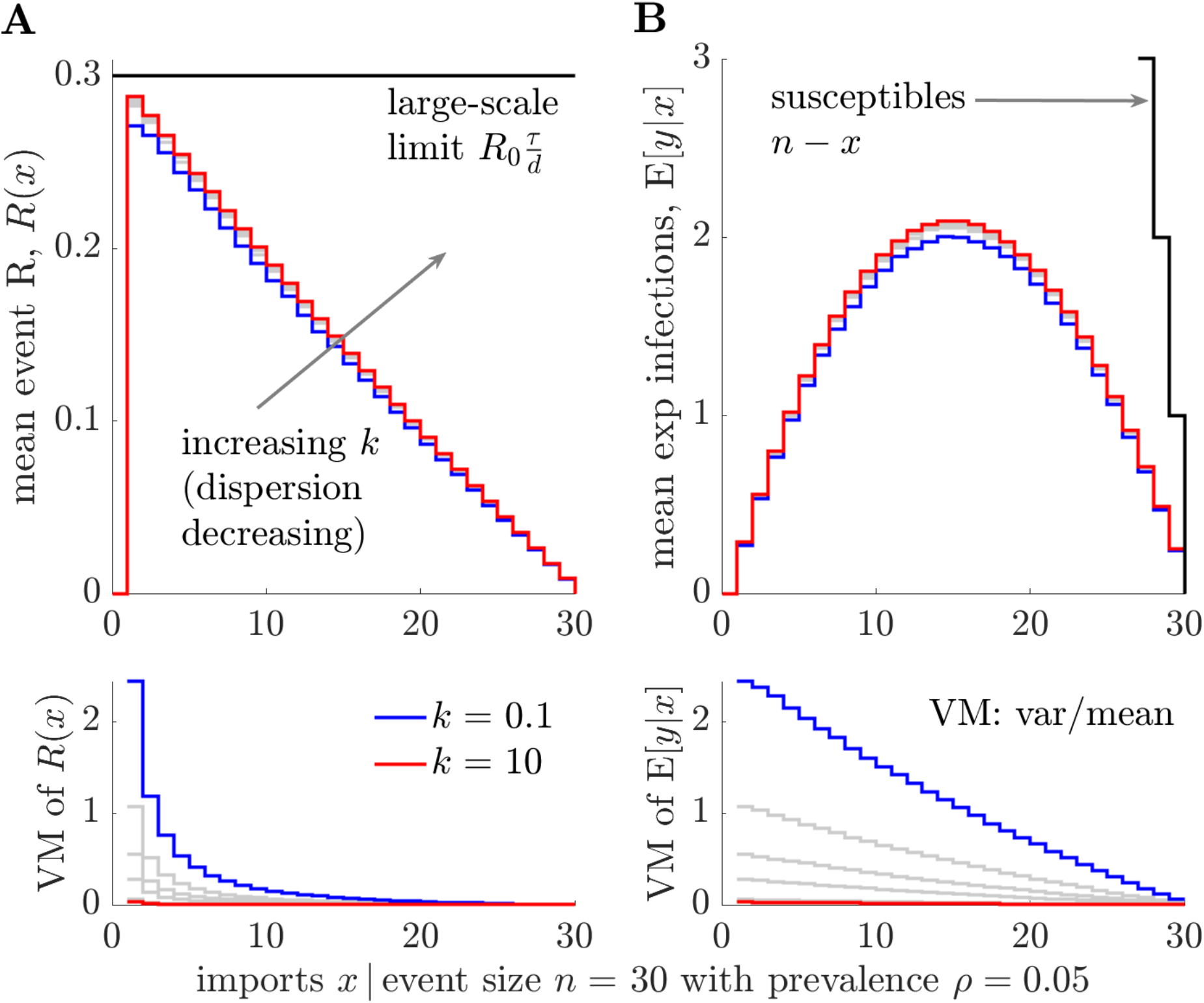
Risk statistics for an event with heterogeneous transmission. We plot the mean (**E**[.], top subfigures) and variance to mean ratio (**VM**[.], bottom subfigures) of the small-scale event reproduction number *R*(*x*) (panel A) and the mean count of new infections **E**[*y*|*x, n*] (panel B) as a function of the number of imports *x*. We compute these via **Eqs. (4)-(6)** and compile statistics over 10^E^ samples from heterogeneous offspring distributions with dispersion parameter *k* ranging from 0.1 to 10 (increasing from blue to red, with grey depicting all intermediate values). For comparison, we show the large-scale reproduction number 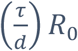 and the number of initial susceptible individuals at the event, *n* − *x*. We repeat this analysis at a larger value of 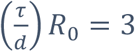 in Supplement **Fig S1**.

Further, increasing heterogeneity (decreasing *k*) increases the variance of our statistics but decreases mean risk as we see from the inversion of the rank of blue to red curves between top and bottom panels in **Fig *2***, with **VM**[.] as the ratio of variance to mean. Last, we see that the dependence of our statistics on the number of imports is substantial and can be as critical as the value of *k* for describing spread. The value of *x* that leads to the largest possible (peak) number of secondary infections at the event is not obvious (and not inferable from *R*(*x*)) as imports both cause infections and reduce the available susceptible individuals. In Supplement **Fig S1** we show how this peak changes and that the mean risk difference can be appreciable in different settings. This underpins the importance of modelling finite event sizes and signifies that a crucial factor driving the risk of acquiring infection at an event is the import distribution **P**(*x*|*n*), which is rarely studied.

### Population prevalence modulates the superspreading potential at events

Having observed the importance of the number of imports, *x* when assessing the transmission risk at events, we explore the influence of the distribution of introductions to the event, **P**(*x*|*n*). Conventionally [25], **P**(*x*|*n*) can be defined as a binomial distribution with the probability of an import being equal to the prevalence of the infection in the wider population, *ρ*. This is our null model, and we explore how it integrates with our proposed event statistics (see **Eqs. (4)-(8)**). We consider epidemics in their initial stages i.e., there is no vaccination- or infection-acquired immunity, so *ρ* is small and there are *n* − *x* susceptible individuals at the event. We maintain parameter settings from **Fig 2** but weight samples of small-scale reproduction numbers and mean numbers of imported infections using **P**(*x*|*n*), which is **Bin**(*x*; *n, ρ*) with *ρ* ranging from 0.01 − 0.1 (1–10%). We compute histograms and statistics of these samples in **Fig 3**.

**Fig 3:**
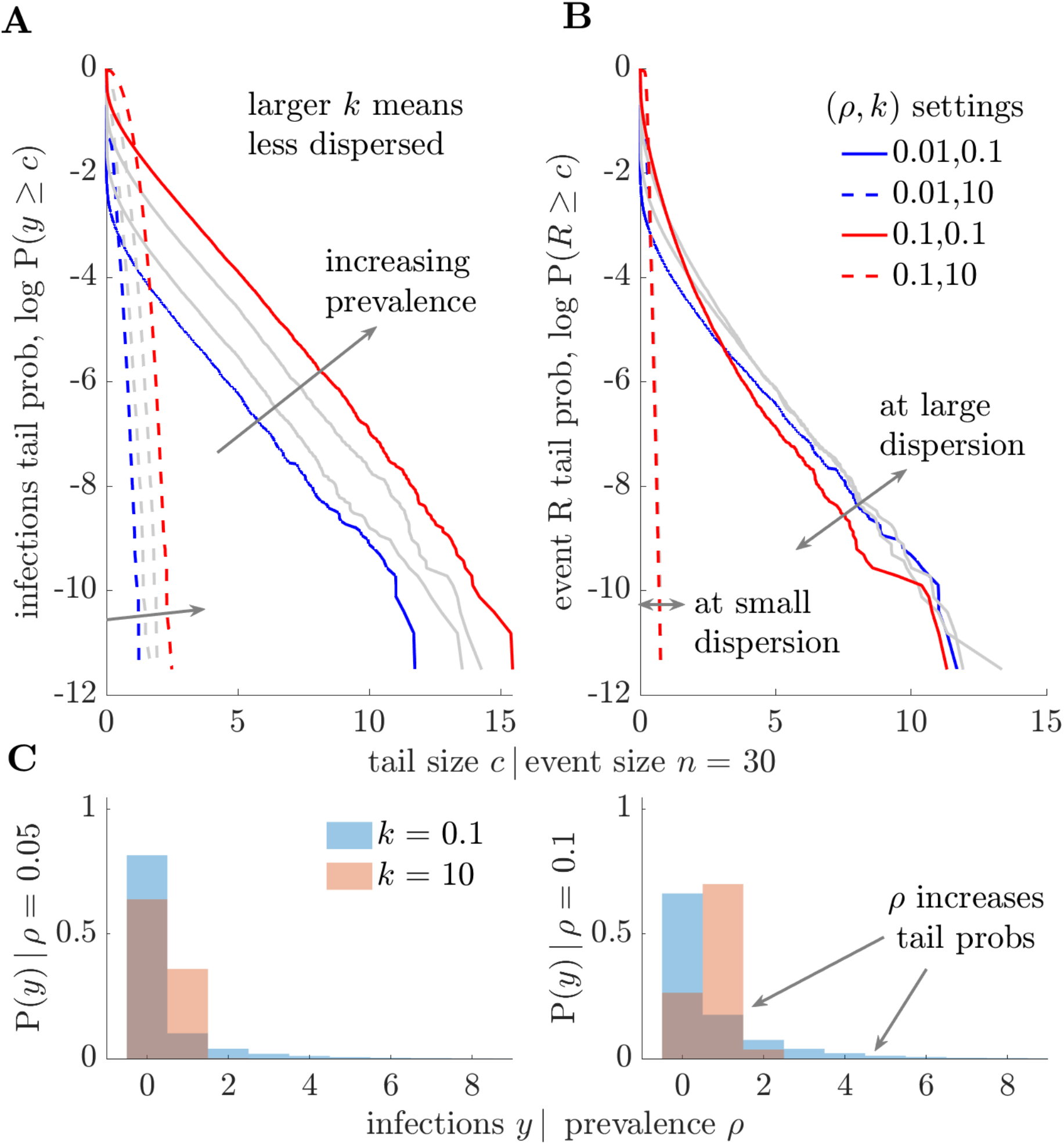
The importation rate magnifies the effects of heterogeneous transmission. We plot the log survival probabilities for the number of new infections *y* (panel A) and related event reproduction numbers *R* (panel B). We account for the probability of *x* imports (distributed as **Bin**(*x*; *n, ρ*)) at an event of size *n* = 30 with the population prevalence as *ρ* (increasing from blue to red with grey indicating intermediate values). Larger **P**(*y* ≥ *c*) signifies more realised heterogeneity (higher likelihoods that disproportionate numbers of infections result from the event), while larger **P**(*R* ≥ *c*) signifies more heterogeneity in transmissibility (higher potential for superspreading events). In panels A-B, dashed curves are for *k* = 10 (spread is mostly homogeneous) and solid curves are for *k* = 0.1 (spread is heterogeneous). We compute these quantities from **Eqs. (4)-(8)**. Panel C shows histograms of 10^E^ samples of *y* at two *ρ* values underpinning panels A-B. Thicker tails or more rightwards mass in these distributions indicate a higher chance of a large number of infections at the event. We repeat this analysis at a larger event size of *n* = 100 in Supplement **Fig S2** for comparison.E

We examine homogeneous (*k* = 10) and heterogeneous (*k* = 0.1) dispersion levels and plot the log survival (or tail) probabilities of realised numbers of new infections *y* and associated small-scale event reproduction numbers *R* in panels A-B of **Fig 3** for different values of *ρ*. We compute these probabilities via **Eqs. (7)-(8)**. Larger values for these probabilities respectively indicate that superspreading is more likely (i.e., disproportionately more infections than **E**_het_ [**E**_imp_[*y*|*n*]] occur) and that imports have increased potential to cause superspreading (i.e., transmissibility above **E**_het_[*R*_imp_]). This distinction is rarely explored because it is less important at population levels, where superspreading models are commonly used [1,26,28]. However, the limiting finite-size effects of events make this distinction crucial. Histograms of samples of the infections at the event for some of the values of *ρ* in A-B are shown in **Fig 3C**.

We find that increasing prevalence ranks the *y* survival curves for both *k* scenarios (panel A) (at a given threshold tail size *c*, probabilities increase with *ρ*) but has limited impact on the *R* curves (panel B). The latter trend is expected as *ρ* does not change transmissibility. However, the fact that prevalence alone can mediate realised superspreading risk is important and, to our knowledge, unexplored. We confirm this with the histograms of panel C, which have thicker tails or at least more rightward probability mass as *ρ* grows (even at large *k*). The variances of the *y* values (not shown) also rise with *ρ*. We show equivalent analyses for a larger sized event (*n* = 100) in Supplement **Fig S2** and recover similar results. The rate at which infections are introduced is therefore critical to assessing the chances of superspreading at event.

The risk of superspreading is a key determinant of whether cases of disease at the beginning of an outbreak will lead to a major epidemic because local infection clusters can propagate forward, snowballing into wider waves of infections. In standard models, the *R* survival curves correlate strongly with those of *y* [1,28]. However, the added variation we see in the *y* curves in **Fig 3** highlights that superspreading risk is above that expected from *R* alone and, further, that chances of stochastic extinction are reduced (the histograms show **P**(*y* = 0) falling with *ρ*). Understanding the interaction between the import rate (determined by the prevalence) and finite event size effects is therefore essential for accurately inferring the risk of superspreading at an event and hence the chance of epidemic establishment. Next, we demonstrate that the realised superspreading risk can further rise if risk awareness affects event attendance.

### Risk awareness controls importation rates and amplifies superspreading risk

We previously assumed that the importation rate into an event was small, constant and equal to the population infection prevalence *ρ*. However, this is unrealistic as event attendance will depend on individual preferences. Data have found that individual perceptions of infection risk can regulate transmission dynamics and that a spectrum of risk appetites exist in a population [13,30,31]. Many models couple behavioural changes to prevalence [10,17] and prevalence elasticity, in which self-protective behaviours vary with prevalence, have been observed. We hypothesise, for a fixed prevalence baseline, that heterogeneity in individual risk perception (i.e., the risk spectrum) may mean that risk-averse individuals avoid larger events where they expect higher chances of becoming infected. Events with large numbers of attendees are then disproportionately likely to be attended by less risk-averse individuals (those with large risk appetites), who have higher chances of introducing infection to the event.

We explore this idea by altering the null model from the above section in which the probability that an event attendee is already infected is *ρ*. We propose a size-biased model where risk appetite or awareness adjusts the event-scale rate of importation based on event size *n*. We realise this using weights that assign a rate *ϵ*(*n*) that scales with *n* (see Methods) but ensures the total infections imported into all events is conserved on average i.e., overall transmission levels are constrained. We consider a set of *m* events, the *i*^th^ of which has size *n*_*i*_. The weight *w*_<_ is set to increase with *n*_*i*_ but satisfies 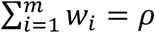. The skew of the *w*_*i*_ i.e., strength of the size-bias, is controlled by the parameter *r*. We apply this model with differing weight strengths *r* using **Eqs. (9)-(10)** and under the parameter settings from **Fig 3**, to obtain **Fig 4**.

**Fig 4:**
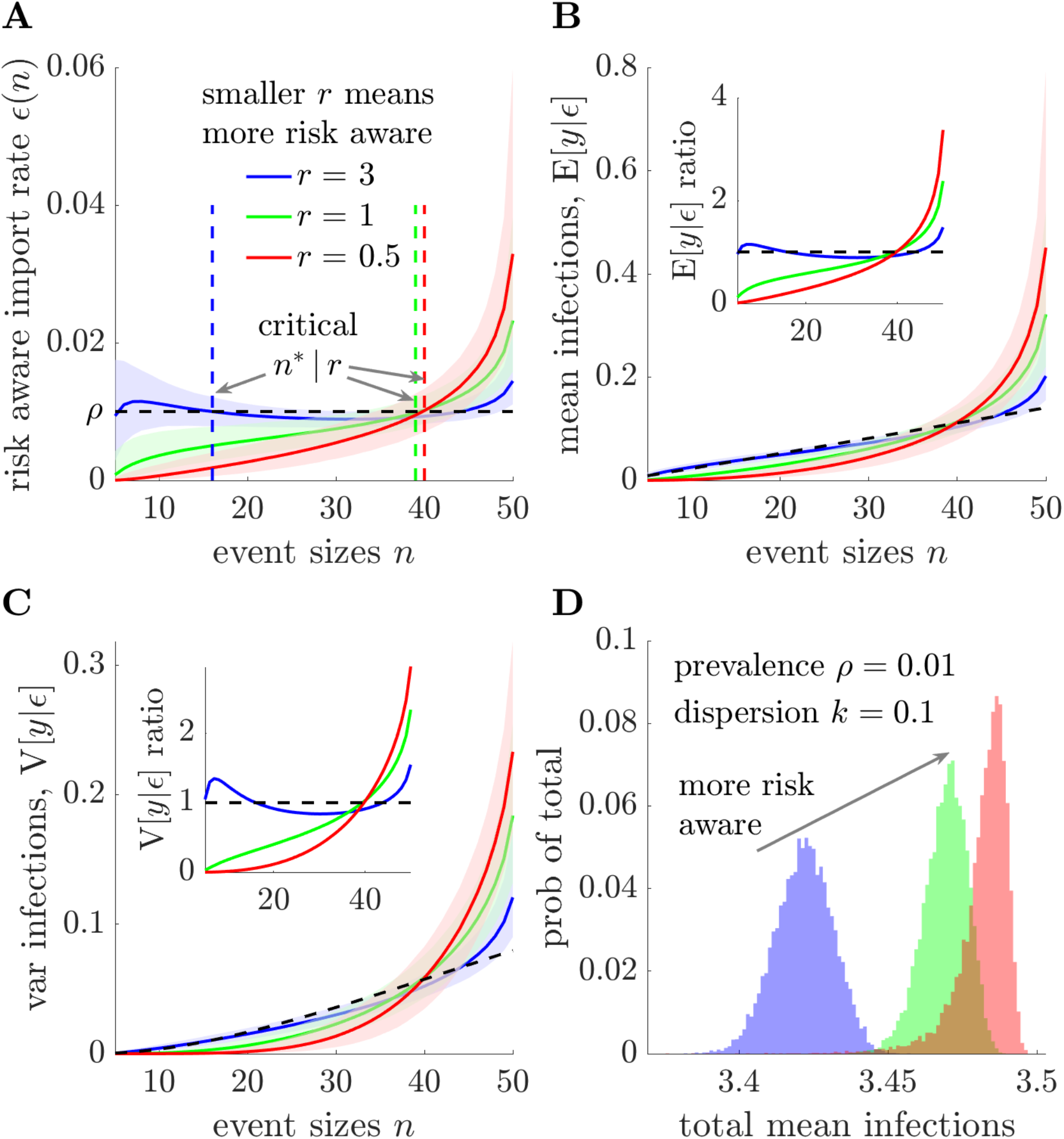
Event size bias substantially elevates the risk of infection. We compare the risk of acquiring infection at an event under models with size-biased import rates due to variability in risk perception against a null model with constant importation rate at the prevalence *ρ*. Panel A shows the size-biased rates *ϵ*(*n*), parametrised by *r*, for *m* = 46 events with sizes spanning E E0. Smaller *r*, decreasing from blue to green to red, indicates more skewed *ϵ*(*n*) functions but conserves the overall infection import rate. The critical event size *n*^*^ demarcates when *ϵ*(*n*) is closest to *ρ* (risk neutral event sizes). Panels B-C illustrate the resulting mean and variance of the number of infections at an event (**E**[*y*|*ϵ*], **V**[*y*|*ϵ*]) relative to the equivalent quantities from the null model (**E**[*y*|*ρ*], **V**[*y*|*ρ*]) for dispersion parameter *k* = 0.1. In panels A-C, we show medians with 95% credible intervals as computed using **Eqs. (7)-(10)**. These marginalise over 10^4^ samples from the distributions of transmission heterogeneity (controlled by *k*) and number of importations (controlled by *ρ* and *ϵ*(*n*)). We also provide ratios of the means of these plots for panels B-C as insets. Panel D shows the total mean number of infections over all events, which remains mostly stable due to our *ϵ*(*n*) constraints.

In **Fig 4**, we study weight choices characterising two risk-awareness levels (green and red), in which the probability that an attendee is an imported infection increases with the event size, a relatively risk-stable case (blue) and a null model (black, dashed) completely neglecting risk-awareness. We show corresponding import rates in panel A of **Fig 4** and compute a critical event size *n*^*^, at which *ϵ*(*n*) is closest to *ρ*. For the risk-aware models (green and red), events above this size have higher infection risk than assumed by conventional (null) models. In panels B-C, we illustrate that size-biasing substantially amplifies the mean and variance of the number of infections *y*, doubling or tripling the risks at larger events, relative to the null model (see insets), for the risk strength parameters and constraints we consider. This amplification outweighs the suppression of infections at smaller events as well as susceptible depletion caused by imports and signifies that risk awareness can strongly shape infection patterns. Our *ϵ*(*n*) constraints limit variations in the total mean infections across events (panel D).

We show the underlying mean, variance and VM ratios of the small-scale, event reproduction numbers as well as VM ratios for infections in Supplement **Fig S3**. These support the trends in **Fig 4**. We also repeat this analysis for epidemics with homogeneous spread (*k* = 10) in Supplement **Fig S4**. Interestingly, while the variance in the number of infections is smaller, the ratios of the means and variances among risk aware and the null model (panels B-C of **Fig S4**) are similar and rise with *n*, indicating that risk awareness alone can introduce additional superspreading risk. These results (with **Fig 3**) mean that neglecting the risk spectrum within host populations, as is done in conventional models where the probability that an attendee is initially infected is set solely by the prevalence *ρ*, can lead to substantial underestimation of the likelihood of superspreading.

We confirm this in **Fig 5**, where we illustrate how log survival or tail probabilities of infections (log **P**(*y* ≥ *c*)) change with the risk awareness strength *r* and *ρ*. In panel A, we fix *r* and find that the median risk of infections, relative to the null risk-neutral model, is largely unchanged. This verifies that the skew of the size-bias from risk awareness is a key variable. In panel B, we see how median relative risk at larger events increases with risk awareness levels (i.e., as *r* falls), for fixed *ρ*. We highlight two event sizes *n* = (24, 48), which have relative risks below and above 1 (dashed lines in A-B) and examine their tail probabilities in panels C-D. In C we find that, for both sizes, superspreading risk rises with prevalence as tail probabilities at any threshold *c* scale with *ρ*. In D we note that risk awareness at a given prevalence can reduce likelihood of superspreading at smaller events but considerably amplify the superspreading risk at larger events (seen as an inversion in the ranking of curves from blue to red).

**Fig 5:**
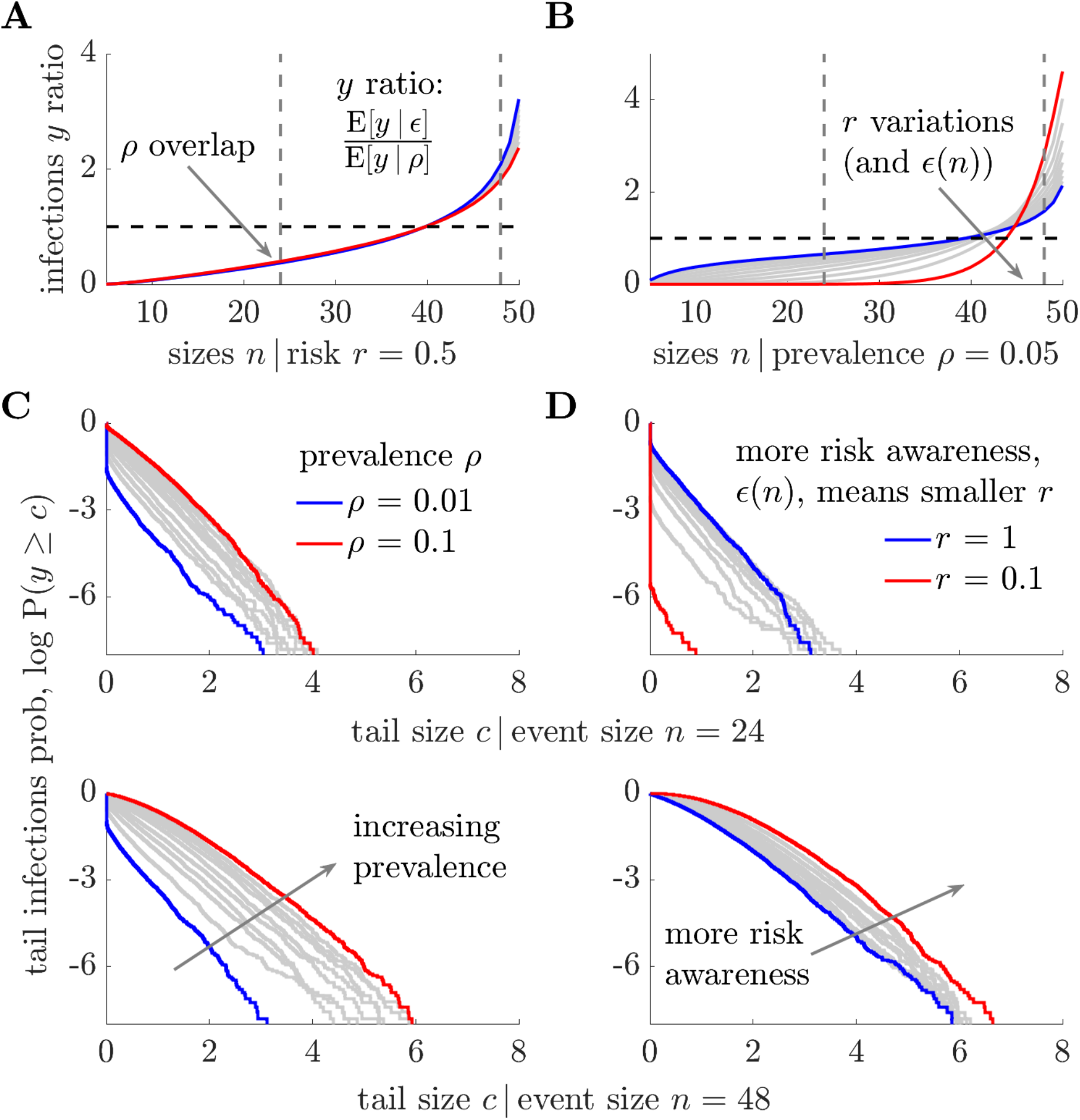
Superspreading risk increases with risk awareness. We repeat the analyses in **Fig 4** but for varying prevalence rates *ρ* at a given risk-awareness strength *r* = 0.E in panel A and for differing strengths at prevalence *ρ* = 0.0E in panel B. These show mean numbers of infections **E**[*y*|E] under risk-aware models relative to that from the null model **E**[*y*|*ρ*] (we plot only medians of distributions for dispersion parameter *k* = 0.1). We demonstrate how risk awareness modulates the risk of superspreading at a medium and large sized events (dashed vertical lines in panels A-B) by exploring tail infection survival probabilities **P**(*y* ≥ *c*) in panels C-D. See Supplement **Fig S3** and **Fig S4** for further accompanying simulations and statistics.

Across our simulations, this amplification from host behaviours can be as much as tenfold (2-2.5 natural log units in the tail probabilities at 4 ≤ *c* ≤ 6 in panel D at *n* = 48). We reinforce these conclusions by computing the associated statistics of new infections (panel A) and our small-scale reproduction numbers (panel B) in **Fig 6**. There we verify that the mean and variance of the number of infections grow with prevalence and the variability in risk awareness within the population, but that the realised heterogeneity is not inferable from the heterogeneity in reproduction numbers. Consequently, the population risk spectrum can independently drive increased superspreading risk at larger events that may have critical ramifications because larger events can support more infections and contribute disproportionately to the establishment of infection in the host population early in an epidemic. As a result, accurate characterisation of small-scale behavioural patterns, in combination with estimation of both the wider-scale prevalence of infection in the population and transmission heterogeneities, are integral to correctly quantifying the risk of superspreading.

**Fig 6:**
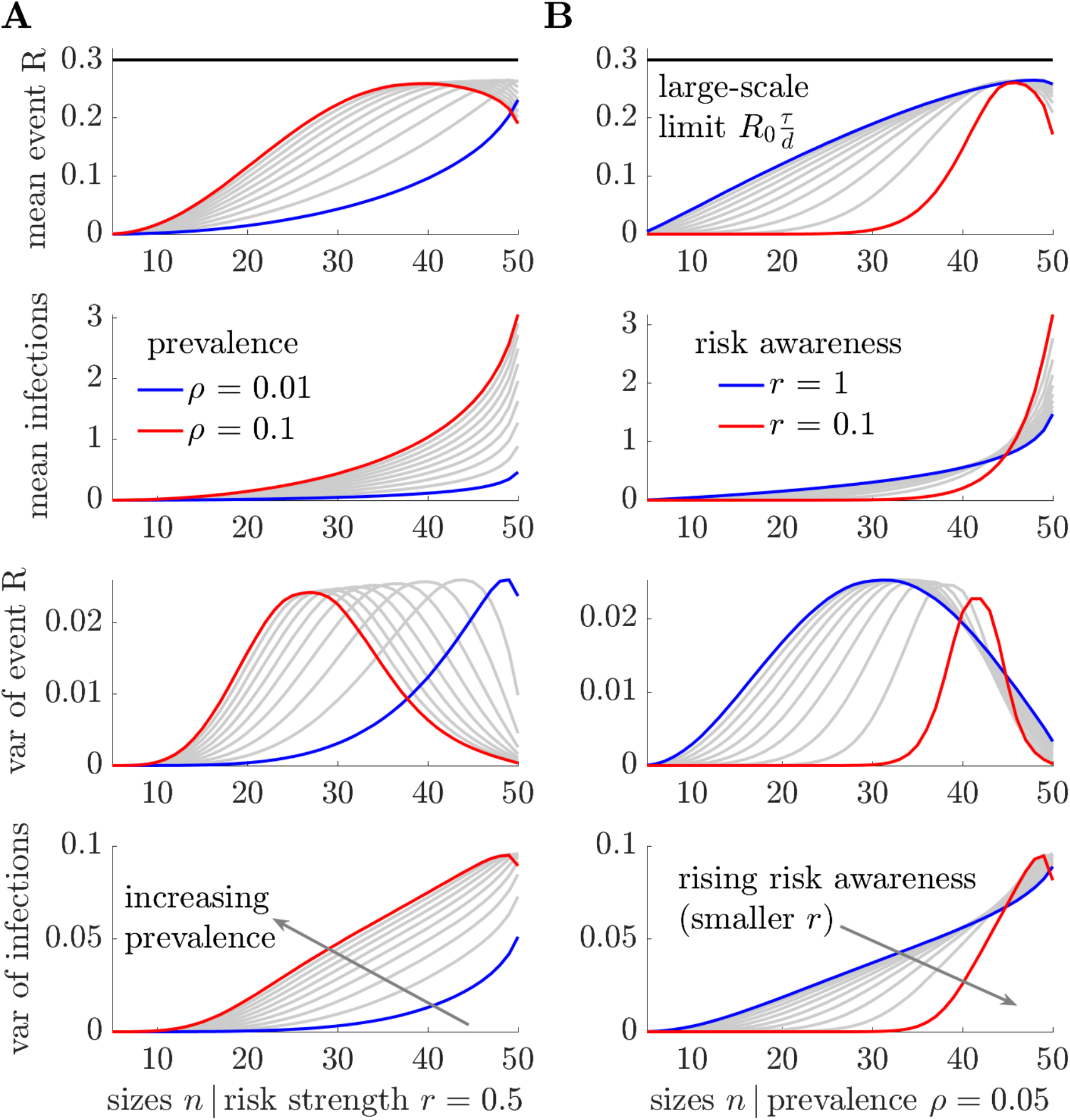
Risk awareness is the key driver of superspreading risk at large events. We expand on the results in **Fig 5** (using the same parameter values) and compute the mean and variance of the small-scale reproduction number *R* and the number of infections at the event *y*. Panel A plots these for differing prevalence *ρ* at fixed risk-awareness strengths *r* (smaller values indicate stronger risk awareness), while B varies *r* at fixed *ρ*. Increasing *ρ* leads to a higher mean number of infections and more variation in the number of infections. Decreasing *r* reduces the mean as well as the variance in the number of infections at smaller events but amplifies them at larger events, increasing the risk of superspreading. The statistics of the reproduction numbers do not reflect the realised numbers of infections (decreases in variances at larger event sizes occur due to saturation) confirming that variability in the risk awareness between individuals is the major driver of the event-level infection patterns.

## Discussion

Human behaviour is a key driver of infectious disease outbreaks, yet it is not often considered in detail in epidemiological modelling studies. While variations in individual perceptions of the risks associated with acquiring infection are known to shape important macroscopic properties of an epidemic, such as disease incidence time series or patterns of spread [11,17,32], few studies have explored how human behaviour impacts the chances of superspreading. This phenomenon, while infrequent and seeded at small scales such as events or gatherings, can generate disproportionate numbers of infections, which can substantially influence large-scale epidemic growth and persistence, particularly during early or emergent stages [3,5,26]. Data or even models connecting the spectrum of infection risk perceptions within host populations to attendance of events are rarely available or studied [9,24]. Here we aspired to resolve this gap by investigating the effects of plausible relationships between human behaviour and event attendance on superspreading and by highlighting how small-scale behavioural data collection can be useful for improving the accuracy of epidemic modelling and hence control.

We developed a computational framework (**Eqs. (1)-(10)**) to model small-scale transmission at events (e.g., weddings, parties, sports matches or concerts) where superspreading may arise and individual behaviour can impact pathogen dynamics. Our framework quantifies, under a standard random mixing assumption, how finite event size effects together with heterogeneities in both the transmissibility among hosts and the rate of introductions of infections to an event, contribute to the numbers of infections generated at that event. This generalises several earlier approaches [23–25] and allowed us to define a within-event (small-scale) reproduction number that measures how importations and individual-level variations impact the transmissibility at events. Our event reproduction number *R*(*x*) meaningfully links to population-level characteristics through its convergence to *R*_0_ when the event size and duration scale asymptotically (see Methods).

Using *R*(*x*), we showed that previous transmissibility metrics, whether derived from branching processes [1] or earlier event-level approaches [24], can overestimate transmission and the number of infections likely to occur at an event (**Fig 2** and Supplement **Fig S1**). This result holds for any model in which the finite supply of susceptible individuals at an event is not accounted for and is exacerbated when there are multiple imported infections, because the number of susceptible individuals that any imported case can infect falls [33]. Further, this finite-size effect highlighted that it is essential to collect data on the number of infections introduced into events to accurately quantify superspreading risk (**Fig 3** and Supplement **Fig S2**), which we found to depend strongly on both the number of imported infections and more conventionally evaluated heterogeneities [3]. This insight hinted at one potential reason why behavioural patterns might affect the likelihood of superspreading – if risk awareness alters the distribution of infections imported into events, then it could also modulate the chances of superspreading occurring at those events.

We explored this possibility using a parsimonious model of human behaviour. We posited that variations in infection risk perceptions or awareness in a population might cause more risk-averse individuals to (probabilistically) avoid larger events, which they believe present a higher risk of acquiring infection. Our framework allowed us to model this as a size-biased weighting on the rate of introducing infections to an event that is a function of both the wider population prevalence and the event size. This draws on real-world observations that there is a spectrum of self-protective behaviours in host populations that are driven by prevalence baselines and heterogeneous risk perceptions [10,12,14,19,20]. Across numerous model simulations, we found, for given event sizes and fixed overall prevalence values, that risk awareness amplifies the chances of superspreading at large events (**Fig 4** and Supplement **Figs S3-4**) but limits transmission at smaller events.

Moreover, as either the prevalence or variability in risk awareness increases (characterised by strength parameter *r*), the chances of superspreading elevate (**Fig 5** and **Fig 6**). This holds irrespective of the inherent heterogeneity in transmissibility at the event (characterised by dispersion parameter *k*), which describes the impact of conventional superspreading drivers such as pathogen biology and host characteristics. Further, the mean, variance and probability of large numbers of infections at the event all support this trend. Since this amplification of within-event transmission occurs precisely at those events with the capacity to support larger numbers of infections (i.e., at larger events, where the effect of susceptible depletion when there are more imports is less), this behavioural mechanism can have major consequences. This may be especially critical during the sensitive, initial stages of potential epidemics, when increased superspreading can spur growth and trigger progression from sporadic outbreaks into sustained waves of infection [3,5].

Although these results underscore the importance of human behaviour in driving infectious disease outbreaks, our approach, like any mathematical modelling study, involved several simplifying assumptions. First, we assumed random mixing within events, so any susceptible individual can interact with any infectious individual with equal probability. In reality, contact networks form at events, and the structure of these networks may differ with the size and type of event. While frameworks for embedding contact structure in epidemiological models exist (for example, multilayer networks can be used to link risk awareness and infection structure [32–34]), they can be complex and difficult to interpret or require high resolution data that are typically unavailable [9,30,35]. We also note that our inclusion of transmission heterogeneities as in [1], together with our weighting of the risk of introductions based on event size, do reflect some features of real-world transmission networks while preserving interpretability.

Second, we assumed that event sizes and durations were pre-determined and fixed the overall import rate across all events. However, risk awareness could itself reduce event durations, sizes, frequencies and thus the prevalence of infections. Conversely, if events are prevented, due to government policy, then less risk-averse individuals may initiate their own unregulated gatherings, which could increase transmission (a rebound effect) [15]. This feedback between behaviour and environment (risk and event properties) might affect chances of superspreading [11]. Characterising this feedback is difficult due to a lack of data linking event properties and human behaviour [9,36]. Future collection and analysis of such data will be vital for grounding hypotheses and avoiding overly strong or prescriptive modelling assumptions.

Third, and relatedly, we did not attempt to model the causes of or temporal changes in the different levels of perceived infection risks among individuals. During initial epidemic stages and especially for novel pathogens, data can be sparse and erratic [35,37]. The perception of the risks associated with acquiring infection may therefore be affected by unreliable reports and major uncertainty about the true risk posed by the invading pathogen. Moreover, these risk perceptions might change across time as population immunity builds or if pharmaceuticals that reduce the severity of infections are introduced at later epidemic stages. It could even be the case that after the epidemic peak, the less risk averse individuals are actually more likely to harbour at least partial immunity. Modelling these poorly understood effects would require strong assumptions that are hard to validate.

To avoid all of the above issues, we only considered initial epidemic stages and focussed on isolating the risk-awareness induced patterns given a set of events and a prevalence baseline. We made a minimal assumption, supported by survey data on population behaviours [12–14], that there is a spectrum of risk about this baseline. Our aim was twofold – to discover how risk perceptions could impact superspreading and to show why collecting auxiliary data describing variations in epidemic-related human behaviour, such as event attendance, are necessary. Improving understanding of the coupling between risk-sensitive behaviours and epidemiology would create a platform for future investigation of the outstanding problems mentioned above.

While our modelling approach is relatively simple, it provides clear evidence that behavioural patterns can substantially amplify the risk of superspreading. Heterogeneity in infection risk perception within host populations, modelled by an event size-biased importation rate *ϵ*(*n*), translates into the potential for substantial transmission at large events during early stages of infectious disease epidemics. Because superspreading plays a pivotal role in epidemic growth and the chance of pathogen establishment, further data are required to uncover and specify the form of *ϵ*(*n*) and the mechanisms that shape the coupling between epidemiological and behavioural patterns. We bolster calls for enhanced surveillance that collects such data [9,30]. Surveys linking perceptions of infection risk with attendance at events [19] are essential to determine when variability in risk awareness may be a principal driver of superspreading. This is important to inform, design and target public health interventions more effectively [4].

## Funding

KVP acknowledges funding from the MRC Centre for Global Infectious Disease Analysis (reference MR/R015600/1), jointly funded by the UK Medical Research Council (MRC) and the UK Foreign, Commonwealth and Development Office (FCDO), under the MRC/FCDO Concordat agreement and is also part of the EDCTP2 programme supported by the European Union. The funders had no role in study design, data collection and analysis, decision to publish, or manuscript preparation. For the purpose of open access, the author has applied a ‘Creative Commons Attribution’ (CC BY) licence to any Author Accepted Manuscript version arising from this submission.

## Data availability statement

All data and code to reproduce the analyses and figures of this work and to compute formulae from the Methods are freely available (in MATLAB) at: https://github.com/kpzoo/smallscaleR.

## Notes

### Competing Interest Statement

The authors have declared no competing interest.

### Summary of Updates

Several analyses have been redone and the associated figures reworked. The context and discussion have also been revised considerably.

