## supplementary information for "Host behaviour driven by awareness of infection risk amplifies the chance of superspreading events"

Kris V Parag[1,2,*] and Robin N Thompson[3]

[1]MRC Centre for Global Infectious Disease Analysis, Imperial College London, London, UK.
[2]NIHR HPRU in Behavioural Science and Evaluation, University of Bristol, Bristol, UK.
[3]Mathematical Institute, University of Oxford, Oxford, UK.

### Auxiliary and supporting analyses

We provide additional simulations supporting the results presented in the main text. In **Fig S1** we repeat the analyses from **Fig 2** but for a larger large-scale reproduction number limit. We observe much more appreciable inversion (the rank of blue to red from heterogeneous to more homogeneous dispersion levels) between the means and variance to mean ratios (VM) of both event reproduction numbers and the numbers of infections occurring due to the event. The skew of the peak in the mean number of infections is now substantial and the finite size effects from the available susceptible individuals to infect (panel B) is more limiting.

In **Fig S2** we repeat the analyses from **Fig 2** and verify that the importation rate of infections at the event remains a key driver of superspreading risk. This point is striking since the tail probabilities for extreme event reproduction numbers decrease with rising prevalence (panel B), whereas the tail probabilities for realised infections at the event increase with prevalence (panel A). This inversion in ranking (from blue to red) suggests that importation rates into a finite event can overpower the expected influence of more traditional drivers of superspreading (in this case heterogeneity in transmissibility, defined by tail reproduction numbers).

Given the observed importance of the importation rate into an event, human behaviour may be expected to play a critical role if it alters that rate. As detailed in the main text, risk averse behaviour can precisely do this, potentially increasing the proportion of attendees at larger events that are already infected (i.e., the import rate). **Figs S3-S4** confirm this intuition. In **Fig 4** of the main text, we demonstrated that risk aversion can substantially elevate likely numbers of new infections at larger events. **Fig S3** verifies that the variance and variance to mean of numbers of infections and small-scale reproduction numbers also increase with risk aversion (for fixed overall transmission levels), increasing the chance of superspreading.

In **Fig S4**, we repeat the simulations from **Fig 4** but consider homogeneous transmission with a dispersion parameter that is 100 times larger. While this causes the variance of the number of infections to be much smaller (panel C), the trends in relative risk and relative variation at different risk awareness strengths remain consistent with **Fig 4**. Elevating risk sensitivity (decreasing the risk aware parameter) consistently and strongly increases the realised risk of superspreading at larger events, beyond some critical event size. The dependence of this risk on import rates into events evidence a need for collecting data on those introductions if we are to accurately measure and respond to the chances of superspreading.

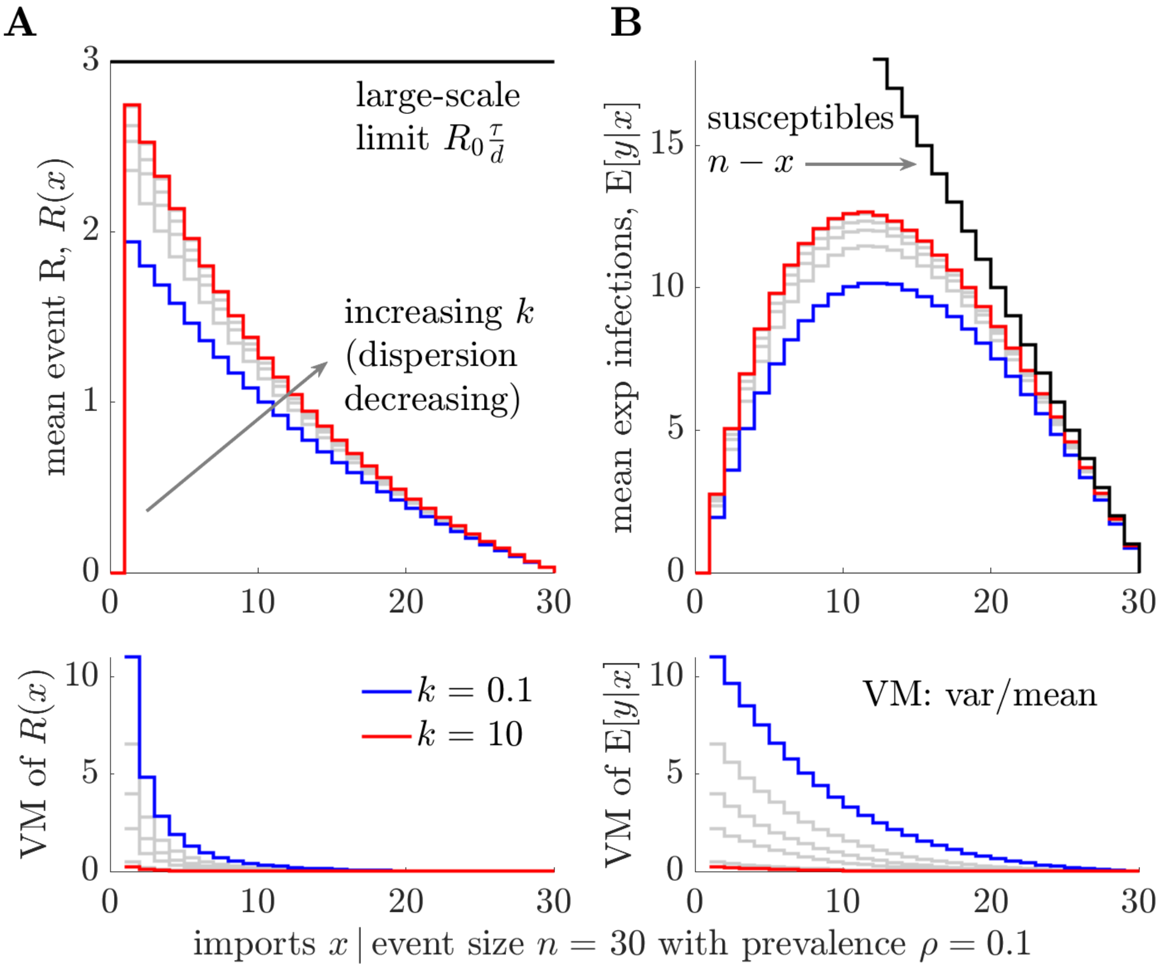


**Fig S1: Risk statistics for an event with heterogeneous transmission.** We replicate **Fig 2** of the main text but at a tenfold larger large-scale reproduction number. We plot the mean (**E**$[.]$, top subfigures) and variance to mean ratio (**VM**$[.]$, bottom subfigures) of the small-scale event reproduction number $R(x)$ (panel A) and the mean count of new infections **E**$[y|x,n]$ (panel B) as a function of the number of imports $x$. We compute these via **Eqs. (4)-(6)** and compile statistics over $10^5$ samples from heterogeneous offspring distributions with dispersion level parameter $k$ ranging from 0.1 to 10 (increasing from blue to red with grey depicting all intermediate values). For comparison, we show the large-scale reproduction number $\left(\frac{\tau}{d}\right) R_0$ and the number of initial susceptible individuals at the event, $n - x$.

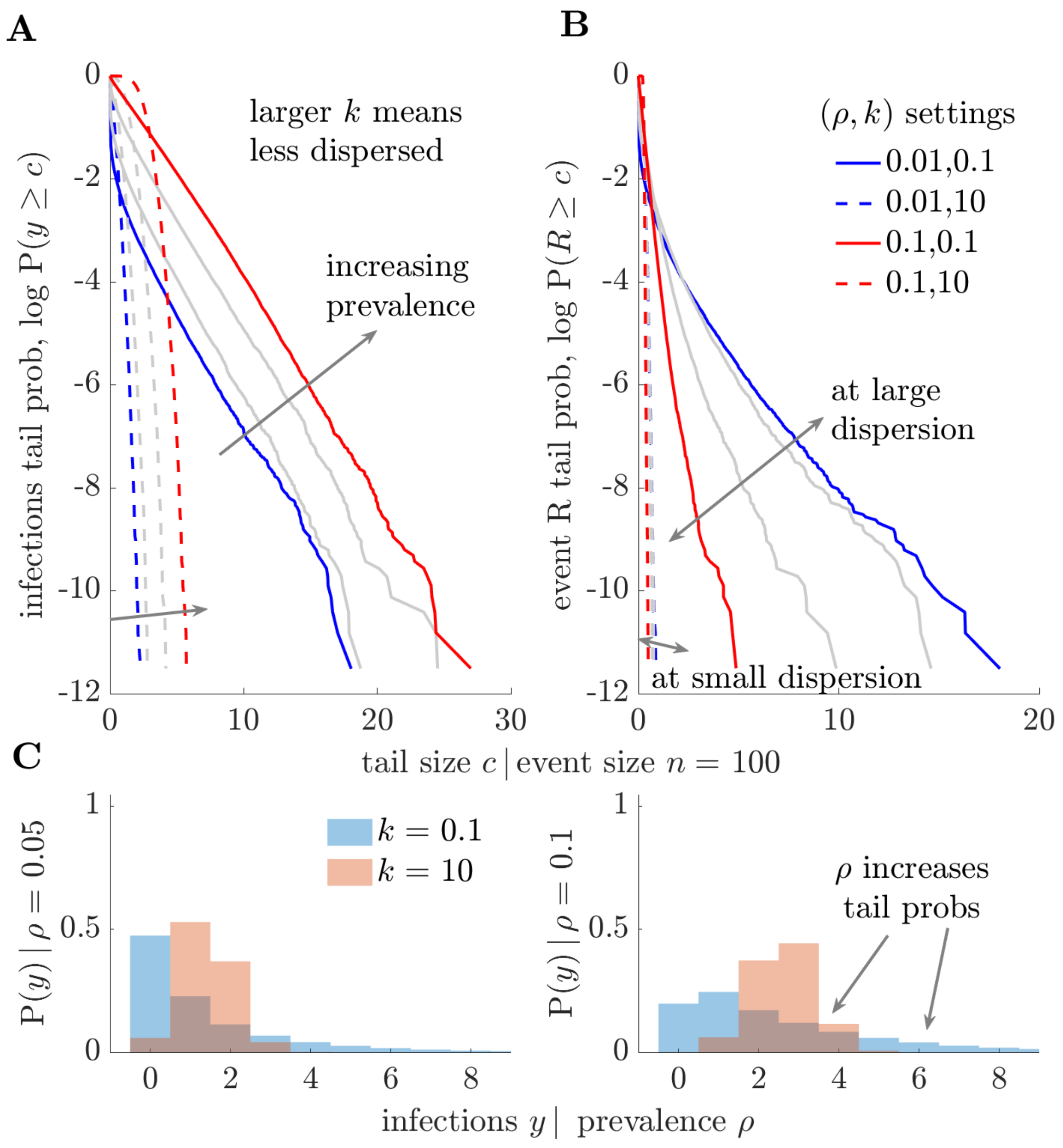


**Fig S2: The importation rate can magnify the chances of superspreading.** We repeat the simulations of **Fig 3** of the main text but for a larger event of size $n = 100$. We plot the log survival probabilities for the number of new infections $y$ (panel A) and the associated event reproduction numbers $R$ (panel B). We account for the probability of $x$ imports (distributed according to **Bin**$(x; n, \rho)$) with the population prevalence as $\rho$ (increasing from blue to red with grey intermediate values). Larger **P**$(y \geq c)$ signifies more realised heterogeneity (higher likelihoods that disproportionate numbers of infections result from the event), while larger

$\mathbf{P}(R \geq c)$ signifies more heterogeneity in transmissibility (higher potential for superspreading events). In panels A-B dashed curves are at $k = 10$ (spread is mostly homogeneous) and solid curves at $k = 0.1$ (spread is heterogeneous). We compute these quantities from **Eqs. (4)-(8)**. Panel C shows histograms of $10^5$ samples of $y$ at two $\rho$ values underpinning panels A-B. Thicker $y$ tails indicate more heterogeneity and occur as $\rho$ increases.

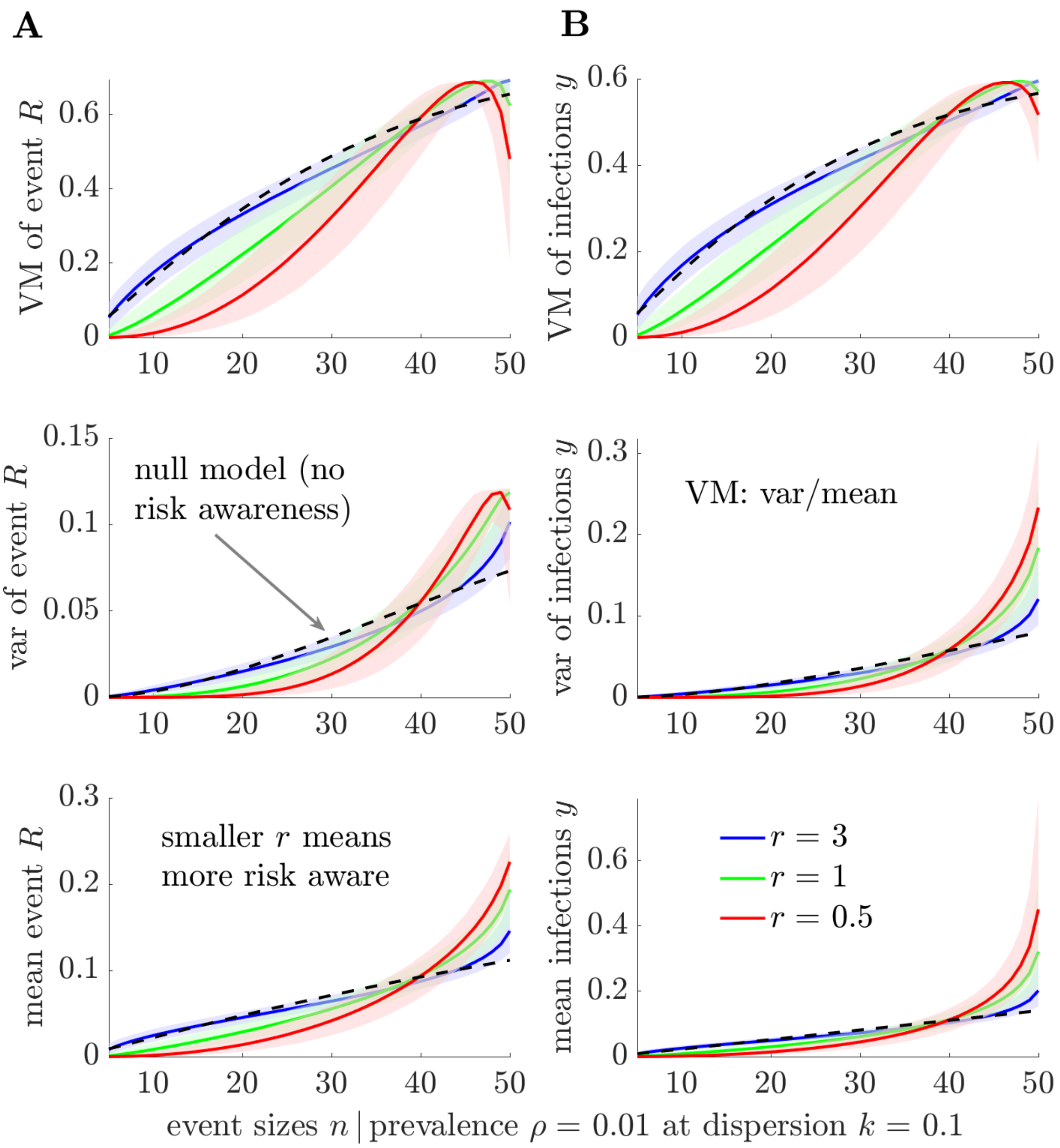


**Fig S3: Risk awareness increases the variability in transmission.** We analyse statistics underpinning **Fig 4** of the main text. We compute variability in the risk of acquiring infection at an event under models with size-biased imports emerging from risk awareness (with a smaller $r$ indicating stronger risk awareness) and compare these to a null model (black dashed) with constant importation rate. We fix the prevalence $\rho$ and the dispersion level $k$. Panel A plots the variance to mean ratios **VM**[.] as well as the underlying variances and means of our small-scale reproduction numbers $R$. Panel B shows equivalent statistics for the new infections $y$ occurring due to the event with those characteristics. All subfigures plot medians with 95% credible intervals, are computed from **Eqs. (7)-(10)** and marginalise over $10^4$ samples from the distributions of transmission heterogeneity and importations.

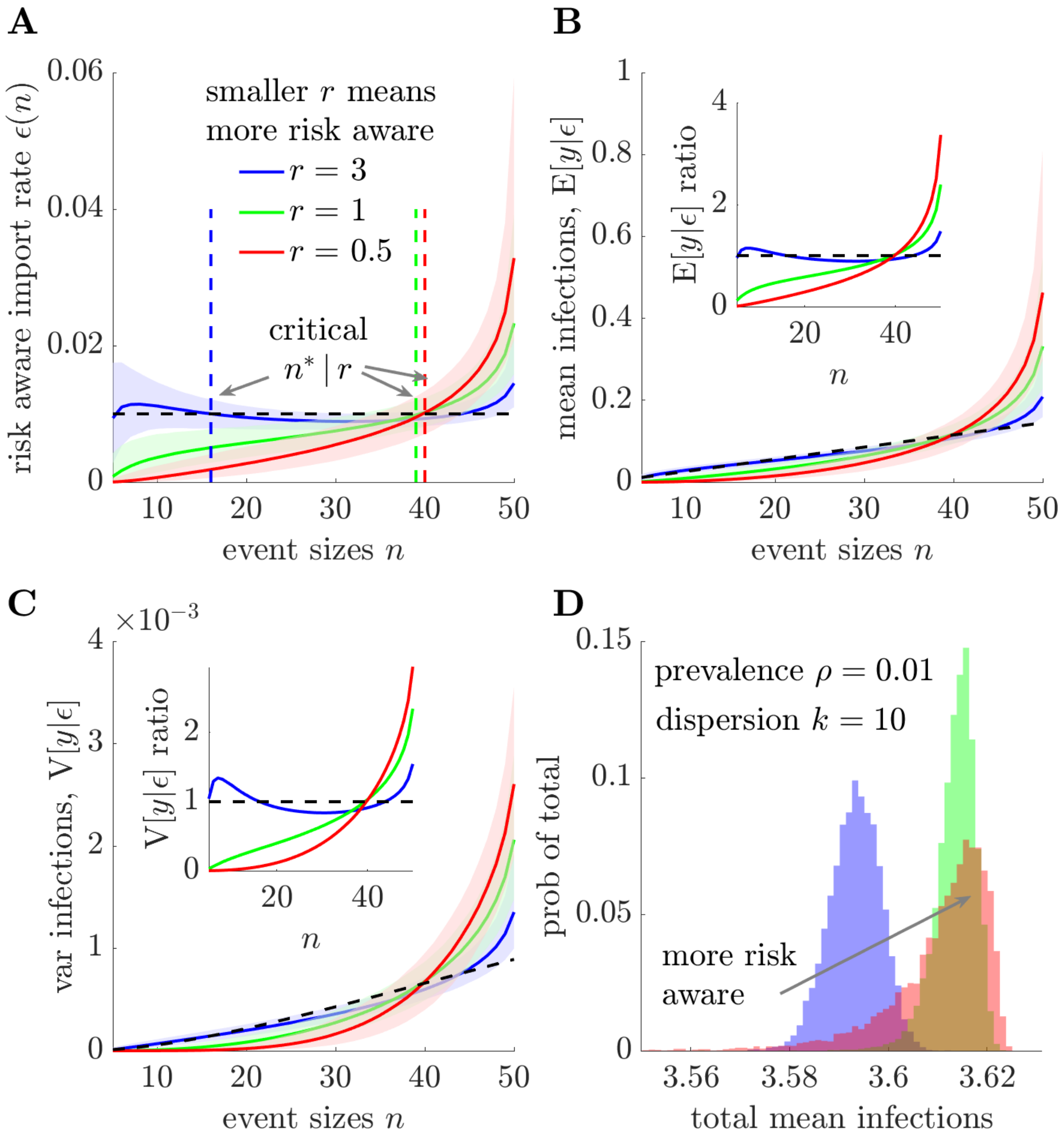


**Fig S4*:* Event size bias substantially elevates the risk of infection at events even under homogeneous transmission.** We repeat the simulations from **Fig 4** of the main text but for more homogeneous transmission by increasing the dispersion parameter by 100 times to $k = 10$. We compare the risk of acquiring infection at an event under models with size-biased introductions emerging from risk awareness to a null model with constant importation rate at the prevalence $\rho$. Panel A shows the size-biased rates $\epsilon(n)$, parametrised by $r$, for $m = 46$ events with sizes spanning $5:50$. Smaller $r$, decreasing from blue to green to red, indicates more skewed $\epsilon(n)$ distributions but conserves the overall transmission level. Critical event size $n^*$ demarcates when $\epsilon(n)$ is closest to $\rho$ (risk neutral event sizes). Panels B-C illustrate the resulting mean and variance of infections at an event (**E**$[y|\epsilon]$, **V**$[y|\epsilon]$) relative to that from the null model (**E**$[y|\rho]$, **V**$[y|\rho]$). Panels A-C plot medians with 95% credible intervals and are computed from **Eqs. (7)-(10)**. These marginalise over $10^4$ samples from the distributions of transmission heterogeneity (controlled by $k$) and importations (controlled by $\rho$ and $\epsilon(n)$). We also provide ratios of the means of these plots for panels B-C as insets. Panel D shows the total mean infections over all events, which is mostly fixed due to constraints on $\epsilon(n)$.

### Data availability

All data and code to reproduce the analyses and figures of this work and to compute formulae from the Methods are freely available (in MATLAB) at: https://github.com/kpzoo/smallscaleR.
